# Mental Health of Keyworkers in the UK during the COVID-19 Pandemic: a Cross-sectional Analysis of a Community Cohort

**DOI:** 10.1101/2020.11.11.20229609

**Authors:** Kieran Ayling, Ru Jia, Trudie Chalder, Adam Massey, Elizabeth Broadbent, Carol Coupland, Kavita Vedhara

## Abstract

**Objectives:** Previous pandemics have resulted in high levels of psychological morbidity among frontline workers. Here we report on the early mental health impact of the COVID-19 pandemic on keyworkers in the UK, as assessed during the first six weeks of nationwide social distancing measures being introduced. Comparisons are made with non-keyworkers, and psychological factors that may be protective to keyworkers’ mental health are explored.

**Design:** Cross-sectional analysis of a community cohort study.

**Methods:** During April 2020, keyworkers (n=1559) and non-keyworkers (n=1436) completed online measures of depression, anxiety, and stress levels as well as explanatory demographic and psychological factors hypothesised to be related to these mental health outcomes.

**Results:** Keyworkers reported significantly higher depression, anxiety, and stress than pre-pandemic population norms. Compared to non-keyworkers, keyworkers were more likely to worry about COVID-19 and perceived they were at higher risk from the virus. This was particularly evident for health and social care keyworkers. Younger keyworkers and those in a clinically increased risk group were more likely to report poorer mental health. Lower positive mood, greater loneliness and worrying more about COVID-19 were all associated with poorer mental health outcomes amongst keyworkers.

**Conclusions:** The mental health impact of the COVID-19 pandemic on keyworkers in the UK has been substantial. Worry about COVID-19 and perceived risk from COVID-19 in keyworkers are understandable given potential increased exposure to the virus. Younger and clinically vulnerable keyworkers may benefit most from any interventions that seek to mitigate the negative mental health impacts of the pandemic.

## Background

In the midst of the first phase of the COVID-19 pandemic, while most of the UK population were required to stay at home, the role of keyworkers stood in stark contrast. Those employed in professions deemed essential (including, but not limited to: health and social care workers, delivery drivers, teachers, and supermarket workers) were expected to continue to work outside of their home, while often shouldering significant additional challenges. Beyond potentially facing greater likelihood of exposure to the SARS CoV-2 virus, many keyworkers have reported increased workloads (YouGov, 2020), shortages of personal protective equipment (British Medical Association Media Team, 2020), and tensions arising between demands of their job and feelings of responsibility to protect immediate family members against infection (McConnell, 2020). Dealing with such conditions presents a potentially significant psychological burden upon keyworkers, with subsequent negative impacts on keyworkers’ mental health.

Worldwide, numerous calls have been made for additional mental health support for keyworkers during the pandemic, particularly those health and social care workers on the frontline (Kinman, Teoh, & Harriss, 2020; Rana, Mukhtar, & Mukhtar, 2020; Sim, 2020; The Lancet, 2020; Xiang et al., 2020). These calls may be well justified given evidence from previous disease outbreaks (such as SARS and MERS) that high prevalence of anxiety, depression, post-traumatic stress disorder, and burnout in keyworkers were observed, both during and after outbreaks (Cabello et al., 2020). Early data from China during the COVID-19 pandemic supports the position that medical healthcare workers had greater insomnia, anxiety and depression compared to non-medical health workers (W. R. Zhang et al., 2020).

While the above highlights the additional risk of psychological morbidity keyworkers (and in particular those in health and social care) may face during the COVID-19 pandemic, conversely, there are aspects of being a keyworker that may be considered psychologically protective. For example, working itself has been shown to be beneficial to mental health (Modini et al., 2016), at least when compared to unemployment. By continuing to go to work, keyworkers may feel a greater sense of continuity and have reduced loneliness compared to non-keyworkers, given the increased number of social interactions they are likely to have. There may also be protective effects from feeling they are helping others (Post, 2011; Schwartz, Meisenhelder, Ma, & Reed, 2003) and greater public appreciation for their contribution. While these factors alone may not be sufficient to completely protect against mental health challenges brought by the pandemic, it is important to understand the factors that contribute to, or mitigate against, worsening mental health among keyworkers at this unprecedented time. This evidence can then inform current and future interventions and/or policy to protect those most in need.

This manuscript seeks to explore the psychological impact of the COVID-19 pandemic on keyworkers during the initial stages of the UK social restrictions that were implemented in late March 2020. Specifically, we explore the following questions: (1) What is the mental health impact of the pandemic on keyworkers? (2) Does the mental health impact of the pandemic differ between keyworkers, keyworker-types, and non-keyworkers?; and (3) what modifiable and non-modifiable factors, are associated with mental health outcomes in keyworkers?

## Methods

### Study Design and Participants

This manuscript presents secondary analyses from the first wave of data collected as part of the COVID Stress and Health Study (Jia et al., 2020). The COVID Stress and Health Study is a longitudinal cohort study examining the psychological and physical effects of the COVID-19 pandemic, and associated social restrictions, on the UK population. The data presented here comes from the first wave of data collection, collected between 3^rd^ April 2020 and 30^th^ April 2020. Ethics and research governance for the study was granted from the University of Nottingham Faculty of Medicine and Health Sciences (ref: 506-2003) and the NHS Health Research Authority (ref: 20/HRA/1858).

Participants were recruited in the community through a social and mainstream media campaign. In addition, to encouraging participation among keyworkers, willing NHS organisations and professional bodies advertised the research through their routine communications. Eligibility criteria stated that participants should be: aged 18 and over; able to give informed consent; able to read English and residing in the UK at the time of completing the survey.

### Procedure & Measures

After consenting to the cohort study, participants completed an online survey implemented through the JISC online survey platform. Validated measures of mental health outcomes included: depression - measured via the 9-item Patient Health Questionnaire (Kroenke, Spitzer, & Williams, 2001; α=0.92); anxiety - measured via the 7-item Generalized Anxiety Disorder Scale (Spitzer, Kroenke, Williams, & Löwe, 2006; α=0.88); and stress - measured via the 4-item Perceived Stress Scale (Cohen & Williamson, 1988; α=0.76).

Demographic factors and potentially modifiable psychological factors hypothesised to be associated with either increased risk of COVID-19 and/or adverse mental health outcomes were also measured. Demographics measured included age, gender, ethnicity, whether the participant was a keyworker and if so what type (e.g., health and social care, teacher etc.), whether or not the participant lived alone, and whether participants were from an identified clinical risk group for COVID-19. Positive mood, which was hypothesised to potentially protective, was measured using the positive affect subscale from the Scale of Positive and Negative Experience (SPANE; Jovanović, Lazić, Gavrilov-Jerković, & Molenaar, 2019) and single items were used to measure perceived loneliness (scale of 1 to 10), worry about contracting COVID-19, perceived risk of getting COVID-19 (scale of 1 to 10), and whether respondents considered they were supporting other people (not including members of immediate family). Question wording for single items can be seen in Supplemental Table 1.

**Table 1:** Cohort Demographics by Keyworker Status

|  | All<br>Keyworkers | Health and<br>social care<br>keyworkers | Non-keyworkers |
| --- | --- | --- | --- |
|  | n (%) | n (%) | n (%) |
| **n** | 1559 (100%) | 1198 (100%) | 1436 (100%) |
| **Gender** |  |  |  |
| Male | 179 (11.5%) | 123 (10.3%) | 271 (18.9%) |
| Female | 1378 (88.4%) | 1073 (89.6%) | 1165 (81.1%) |
| Prefer not to say | 2 (0.1%) | 2 (0.2%) | 0 (0%) |
| **Mean age (SD)** | 43.8 (11.9) | 44.0 (11.6) | 45.4 (17.6) |
| **Age groups (years)** |  |  |  |
| 18-24 | 93 (6.0%) | 56 (4.7%) | 271 (18.9%) |
| 25-34 | 300 (19.3%) | 243 (20.3%) | 228 (15.9%) |
| 35-44 | 376 (24.1%) | 285 (23.8%) | 261 (18.2%) |
| 45-54 | 451 (29.0%) | 353 (29.5%) | 239 (16.6%) |
| 55-64 | 309 (19.8%) | 241 (20.1%) | 261 (18.2%) |
| 65-71 | 29 (1.9%) | 19 (1.6%) | 176 (12.3%) |
| **Ethnicity** |  |  |  |
| White – British, Irish, other | 1405 (90.1%) | 1072 (89.5%) | 1291 (89.9%) |
| Asian/Asian British – Indian, Pakistani, Bangladeshi, other | 66 (4.2%) | 53 (4.4%) | 53 (3.7%) |
| Black/Black British – Caribbean, African, other | 31 (2.0%) | 26 (2.2%) | 11 (0.8%) |
| Chinese/Chinese British | 8 (0.5%) | 8 (0.7%) | 20 (1.4%) |
| Mixed race – White and Black/Black British | 11 (0.7%) | 9 (0.8%) | 8 (0.6%) |
| Middle Eastern/Middle Eastern British – Arab, Turkish, other | 5 (0.3%) | 2 (0.2%) | 18 (1.3%) |
| Mixed race – other | 20 (1.3%) | 17 (1.4%) | 20 (1.3%) |
| Other ethnic group | 11 (0.7%) | 10 (0.8%) | 14 (1.0%) |
| Prefer not to say | 2 (0.1%) | 1 (0.1%) | 3 (0.1%) |
| **Relationship status** |  |  |  |
| Single, never married | 234 (15.0%) | 166 (13.9%) | 338 (23.5%) |
| Single, divorced or widowed | 116 (7.4%) | 94 (7.9%) | 113 (7.9%) |
| In a relationship/married but living apart | 120 (7.7%) | 87 (7.3%) | 134 (9.3%) |
| In a relationship/married and cohabiting | 1077 (69.1%) | 840 (70.1%) | 840 (58.5%) |
| Prefer not to say | 12 (0.8%) | 11 (0.9%) | 11 (0.8%) |
| **Education (highest level of attainment)** |  |  |  |
| No qualifications | 11 (0.7%) | 7 (0.6%) | 14 (1.0%) |
| Completed GSCE/CSE/O-levels or equivalent | 153 (9.8%) | 105 (8.8%) | 85 (5.9%) |
| Completed post-16 vocational course | 54 (3.5%) | 34 (2.8%) | 41 (2.9%) |
| A-levels or equivalent (at school until aged 18) | 156 (10.0%) | 99 (8.3%) | 238 (16.6%) |
| Undergraduate degree or professional qualification | 665 (42.7%) | 532 (44.4%) | 598 (41.6%) |
| Postgraduate degree | 505 (32.4%) | 413 (34.5%) | 450 (31.3%) |
| Prefer not to say | 15 (1.0%) | 8 (0.7%) | 10 (0.7%) |
| **Place of residence** |  |  |  |
| South West England | 104 (6.7%) | 73 (6.1%) | 123 (8.6%) |
| East Midlands | 242 (15.5%) | 152 (12.7%) | 487 (33.9%) |
| Yorkshire and Humber | 176 (11.3%) | 144 (12.0%) | 110 (7.7%) |
| North East | 50 (3.2%) | 26 (2.2%) | 95 (6.6%) |
| East of England | 79 (5.1%) | 61 (5.1%) | 69 (4.8%) |
| North West | 249 (16.0%) | 202 (16.9%) | 102 (7.1%) |
| South East England | 216 (13.9%) | 165 (13.8%) | 174 (12.1%) |
| Greater London | 199 (12.8%) | 166 (13.9%) | 123 (8.6%) |
| West Midlands | 90 (5.8%) | 73 (6.1%) | 75 (5.2%) |
| Northern Ireland | 1 (0.1%) | 1 (0.1%) | 7 (0.5%) |
| Wales | 38 (2.4%) | 31 (3.0%) | 33 (2.3%) |
| Scotland | 115 (7.4%) | 104 (8.7%) | 38 (2.7%) |
| **Keyworker status** |  |  |  |
| Health, social care or relevant related support worker | 1198 (76.8%) | 1198 (100%) | N/A |
| Teacher or childcare worker still travelling in to work | 70 (4.5%) | N/A | N/A |
| Transport worker still travelling in to work | 1 (0.1%) | N/A | N/A |
| Food chain worker (e.g. production, sale, delivery) | 33 (2.1%) | N/A | N/A |
| Key public services worker (e.g. justice staff, religious staff, public service journalist or mortuary worker) | 22 (1.4%) | N/A | N/A |
| Local or national government worker delivering essential public services | 41 (2.6%) | N/A | N/A |
| Utility worker (e.g. energy, sewerage, postal service) | 5 (0.3%) | N/A | N/A |
| Public safety or national security worker | 11 (0.7%) | N/A | N/A |
| Worker involved in medicines or protective equipment production or distribution | 10 (0.6%) | N/A | N/A |
| Other 'key worker' role not listed | 168 (10.8%) | N/A | N/A |
| **Living alone** |  |  |  |
| Living alone | 182 (11.7%) | 147 (12.3%) | 189 (13.2%) |
| Living with others | 1377 (88.3%) | 1051 (87.7%) | 1247 (86.8%) |
| **Covid-19 risk status** |  |  |  |
| Most at risk (e.g. suffering from advanced cancer, severe asthma/COPD, etc.) | 61 (3.9%) | 42 (3.5%) | 56 (3.9%) |
| At increased risk (e.g., being pregnant, aged over 70, etc.) | 208 (13.3%) | 163 (13.6%) | 225 (15.7%) |
| Not at-risk | 1290 (82.8%) | 993 (82.9%) | 1155 (80.4%) |
| **Supporting Others (outside of immediate family)** | 1084 (69.5%) | 841 (70.2%) | 757 (49.2%) |

### Statistical analysis

Statistical analyses were performed using STATA (version 16). As the oldest keyworker was aged 71, we restricted all analyses to only include those aged less than 72 years. We first summarised psychological outcomes and participant characteristics in keyworkers and non-keyworkers with appropriate summary statistics. For depression and anxiety, we additionally examined numbers and percentages of keyworkers and non-keyworkers scoring 10 or greater (indicating moderate or severe levels). This is the threshold used to determine access to high intensity psychological therapies in Improving Access to Psychological Therapies (IAPT) services in the NHS.

To compare levels of psychological morbidity among keyworkers to pre-pandemic normative values we conducted independent samples t-tests against published normative population data (depression: Kocalevent, Hinz, & Brähler, 2013; anxiety: Löwe et al., 2008; stress: Warttig, Forshaw, South, & White, 2013). Additional comparisons were made by splitting the cohort by keyworker type (health and social care keyworkers vs other keyworkers), gender, and age group and compared against matched normative data where available.

To examine whether there were differences between keyworkers and non-keyworkers on the psychological factors measured we performed univariable and multivariable linear regression analyses, controlling for age, gender (male/female), ethnicity (white/BAME), clinical risk group status (“not at risk”/”increased risk”/”most at risk”) and whether they lived alone. Based on examination of descriptive statistics, further analyses were performed to examine possible interactions between gender and keyworker status. Assumptions of linear regression (normality and homoscedasticity of residuals, linearity with continuous variables) and presence of outliers were assessed graphically. Multicollinearity was assessed using variance inflation factors. Square root transformations were used for depression and anxiety scores to satisfy assumptions. Worry about COVID-19 was treated as a categorical variable in all models, with “occasional worry” treated as the reference value as this was the most common response. Multinomial logistic regression analyses were used when considering worry about COVID-19 as a dependent variable comparing relative risk ratios against “occasional worry”. We repeated these analyses in order to examine health and social care keyworkers and other keyworkers separately.

To explore factors, both modifiable and non-modifiable, that may be associated with stress, anxiety and depression among all keyworkers, we first examined associations between demographic factors and these mental health outcomes using multivariable linear regression analyses as described above, before adding potentially modifiable psychological and behavioural factors (positive mood, loneliness, worry about COVID-19, supporting others outside of immediate family) to the models.

## Results

In total, 3097 eligible individuals participated in the study, of whom 2995 were aged 71 or younger. Of these, 1559 (52.1%) self-identified as a keyworker. Health, social-care or relevant related support workers were the largest category of keyworkers accounting for 76.8% of all keyworker participants. Characteristics of keyworker and non-keyworker participants are presented in Table 1. While characteristics were broadly comparable between keyworkers and non-keyworkers, there were more non-keyworkers in the 65-71 age group (12.3% of non-keyworker respondents compared to 1.9% of keyworker respondents). Keyworkers were also less likely to report being “single, never married” than non-keyworkers (15.0% vs 23.5%) and more likely to consider themselves as supporting others outside of immediate family (69.5% vs 49.2%).

### What is the mental health impact of the pandemic on keyworkers?

Keyworkers reported significantly higher levels of stress, anxiety, and depression than pre-pandemic normative values, with highest levels evident in women and younger respondents (see Table 2). These findings were also true for non-keyworkers. The only notable exception to this pattern was for stress scores, which were significantly higher among male keyworkers, compared with pre-pandemic population norms (all male keyworkers mean: 6.37 vs 5.56; *p* = .003). In contrast stress scores in female keyworkers did not differ significantly from pre-pandemic population norms (all female keyworkers mean: 6.43 vs pre-pandemic norm: 6.38, *p* = 0.71), although female non-keyworkers had significantly higher stress scores than pre-pandemic population norms (mean: 6.87 vs pre-pandemic norm 6.38, *p* = .001).

**Table 2:** Depression (PHQ-9), anxiety (GAD-7) and stress (PSS-4) scores a for all keyworkers, health and social care keyworkers alone and non-keyworkers

|  | DEPRESSION score |  |  |  | ANXIETY score |  |  |  | STRESS score |  |  |  |
| --- | --- | --- | --- | --- | --- | --- | --- | --- | --- | --- | --- | --- |
|  | All keyworkers | Health and social care worker | Non-keyworker | <sup>a</sup>Norms | All keyworkers | Health and social care worker | Non-keyworker | <sup>a</sup>Norms | All keyworkers | Health and social care worker | Non-keyworker | <sup>a</sup>Norms |
|  | Mean (SD) | Mean (SD) | Mean (SD) | Mean (SD) | Mean (SD) | Mean (SD) | Mean (SD) | Mean (SD) | Mean (SD) | Mean (SD) | Mean (SD) | Mean (SD) |
| **Total Score** | 7.90 (5.9)*** | 7.76 (5.8)*** | 7.74 (6.2)*** | 2.91 (3.5) | 6.85 (5.5)*** | 6.74 (5.4)*** | 6.58 (5.6)*** | 2.95 (3.4) | 6.42 (3.2)** | 6.30 (3.2) | 6.65 (3.4)*** | 6.11 (3.1) |
| **Gender** |  |  |  |  |  |  |  |  |  |  |  |  |
| Male | 7.34 (6.2)*** | 7.23 (5.9)*** | 6.31 (6.0)*** | 2.7 (3.5) | 6.19 (5.7)*** | 6.17 (5.7)*** | 4.91 (5.3)*** | 2.66 (3.2) | 6.37 (3.3)** | 6.21 (3.3)* | 5.70 (3.3) | 5.56 (3.0) |
| Female | 7.98 (5.9)*** | 7.84 (5.8)*** | 8.07 (6.2)*** | 3.1 (3.5) | 6.95 (5.5)*** | 6.81 (5.4)*** | 6.97 (5.6)*** | 3.20 (3.5) | 6.43 (3.2) | 6.32 (3.1) | 6.87 (3.4)*** | 6.38 (3.2) |
| **Age groups (years)** |  |  |  |  |  |  |  |  |  |  |  |  |
| 18-24 | 10.83 (6.5) | 11.05 (6.3) | 11.38 (6.3) | .. | 9.34 (6.1) | 9.93 (6.3) | 8.91 (5.9) | .. | 7.89 (3.1) | 7.96 (2.8) | 8.21 (3.4) | .. |
| 25-34 | 8.83 (5.8)*** | 8.71 (5.8)*** | 8.63 (6.1)*** | 2.3 (3.2) | 8.01 (5.5)*** | 7.94 (5.6)*** | 7.36 (5.7)*** | 2.81 (3.3) | 6.97 (3.2) | 6.79 (3.2) | 6.90 (3.5) | .. |
| 35-44 | 8.46 (6.0)*** | 8.21 (5.7)*** | 7.90 (5.9)*** | 2.6 (3.5) | 7.19 (5.5)*** | 7.09 (5.3)*** | 7.33 (5.8)*** | 2.82 (3.3) | 6.51 (3.2) | 6.28 (3.1) | 6.91 (3.2) | .. |
| 45-54 | 7.37 (5.6)*** | 7.33 (5.6)*** | 7.23 (5.9)*** | 2.8 (3.5) | 6.41 (5.3)*** | 6.32 (5.2)*** | 6.04 (5.2)*** | 3.14 (3.4) | 6.14 (3.1) | 6.12 (3.1) | 6.20 (3.0) | .. |
| 55-64 | 6.52 (5.5)*** | 6.51 (5.4)*** | 6.15 (5.7)*** | 3.2 (3.5) | 5.49 (5.2)*** | 5.28 (4.9)*** | 5.37 (5.1)*** | 3.25 (3.6) | 5.91 (3.1) | 5.87 (3.1) | 5.96 (3.3) | .. |
| 65-71 | 4.69 (6.0) | 3.68 (4.9) | 3.79 (4.1) | .. | 4.24 (5.6) | 3.37 (5.4) | 3.39 (3.6) | .. | 4.66 (3.2) | 4.32 (3.1) | 5.21 (3.0) | .. |

PHQ-9, the 9-item Patient Health Questionnaire (Kroenke et al., 2001); GAD-7, the 7-item Generalized Anxiety Disorder Scale (Spitzer et al., 2006); PSS-4, the 4-item Perceived Stress Scale (Cohen et al., 1988).
<sup>a</sup>Independent sample t-tests were used for comparisons between mean values in the cohort and previously reported social normative data (depression, Kocalevent et al., 2013; anxiety, Löwe et al., 2008; stress, Warttig et al., 2013). No comparative normative data was available for stress by age groups, or the 18-24 and 65-71 age groups for any outcome.
\*\*\*P<0.001, \*\*P<0.01, \*P<0.05

We observed that 66% of all keyworkers (and 66% of health and social care keyworkers) reported symptoms of depression (score≥ 5), and that the proportion meeting criteria for high intensity support according to NHS criteria (score ≥ 10) was 33% of keyworkers (and 32% of health and social care keyworkers). These proportions are consistent with those observed for non-keyworkers, where 63% reported symptoms of depression and 32% met criteria for high intensity support (see Table 3). Very similar findings were evident for anxiety, with 59% of all keyworkers (and 59% of health and social care keyworkers) reporting symptoms of anxiety (score≥ 5). 27% of all key workers met published criteria for high intensity support (score≥ 10). Findings were again consistent with the non-keyworker group, 56% of whom reported symptoms of anxiety and 26% of whom met criteria for high intensity support.

**Table 3:** Prevalence of depression and anxiety cases among all keyworkers, health and social keyworker subgroup and non-keyworkers

|  |  | All keyworkers |  | Health and social care workers |  | Non-keyworkers |  |
| --- | --- | --- | --- | --- | --- | --- | --- |
|  | Categories | n | % | n | % | n | % |
| **Depression (PHQ-9<sup>b</sup>)** | *No-Minimal Depression (0-4)* | 526 | 33.7 | 406 | 33.9 | 530 | 36.9 |
|  | *Mild Depression (5-9)* | 516 | 33.1 | 408 | 34.1 | 452 | 31.5 |
|  | *Moderate Depression (10-14)* | 286 | 18.4 | 215 | 18.0 | 237 | 16.5 |
|  | *Moderately Severe Depression (15-19)* | 150 | 9.6 | 112 | 9.4 | 123 | 8.6 |
|  | *Severe Depression (20-27)* | 81 | 5.2 | 57 | 4.8 | 94 | 6.6 |
| **Anxiety (GAD-7<sup>b</sup>)** | *No-Minimal Anxiety (0-4)* | 638 | 40.9 | 496 | 41.4 | 630 | 43.9 |
|  | *Mild Anxiety (5-9)* | 493 | 31.6 | 394 | 32.1 | 433 | 30.2 |
|  | *Moderate Anxiety (10-14)* | 238 | 15.3 | 182 | 15.2 | 189 | 13.2 |
|  | *Severe Anxiety (15-21)* | 190 | 12.2 | 136 | 11.4 | 184 | 12.8 |

<sup>a</sup> Cut-offs for categories in line with published guidelines for PHQ-9 and GAD-7.
<sup>b</sup> PHQ-9, the 9-item Patient Health Questionnaire (Kroenke et al., 2001); GAD-7, the 7-item Generalized Anxiety Disorder Scale (Spitzer et al., 2006).

### Does the mental health impact of the pandemic differ between keyworkers, keyworker-types, and non-keyworkers?

Mean scores for psychological variables split by keyworker status can be seen in Tables 2, 3 and 4. Regression analyses were first performed to examine whether there were differences in psychological variables between all keyworkers and non-keyworkers (results of unadjusted and adjusted models can be seen in the supplemental Tables 2-25). Considering all keyworkers, after controlling for age, gender, whether the participant lived alone, ethnicity, and clinical risk group status we observed that keyworkers reported significantly lower levels of stress (B = -0.24, 95% CI: [-0.47, -0.01], *p* = 0.037) but also reported greater perceived risk of getting COVID-19 (B = 1.48, 95% CI: [1.32, 1.65], *p* < .001) compared to non-keyworkers. Additionally, keyworkers were more likely to spend “much of their time” worrying about COVID-19, rather than just “occasionally”, (Relative Risk Ratio = 1.39, 95% CI: [1.11, 1.73], *p* = 0.004), and relatedly were less likely to “not worry” about COVID-19 compared with non-keyworkers (Relative Risk Ratio = 0.75, 95% CI: [0.61, 0.92], *p =* 0.006). No statistically significant differences between keyworkers and non-keyworkers were evident for other psychological variables including depression, anxiety, loneliness, or positive mood.

**Table 4:** Descriptive data for modifiable psychological characteristics by keyworker status

|  | Keyworkers |  |  |
| --- | --- | --- | --- |
|  | All keyworkers | Health and social care workers | Non-keyworkers |
| **Perceived loneliness** |  |  |  |
| Mean (SD) | 3.80 (2.7) | 3.68 (2.7) | 4.00 (2.8) |
| **Positive mood** |  |  |  |
| Mean (SD) | 18.9 (5.0) | 19.0 (4.9) | 18.88 (5.1) |
| **Perceived risk<sup>a</sup>** |  |  |  |
| Mean (SD) | 5.55 (2.2) | 5.7 (2.2) | 4.07 (2.0) |
| **Worry about getting Covid-19** |  |  |  |
| No worry (n, %) | 215 (13.8%) | 158 (13.2) | 277 (19.3%) |
| Occasional worry (n, %) | 1027 (65.9%) | 804 (67.1%) | 947 (66.0%) |
| Much worry (n, %) | 244 (15.7%) | 180 (15.0%) | 164 (11.4%) |
| Most worry (n, %) | 73 (4.7%) | 56 (4.7%) | 48 (3.4%) |

<sup>a</sup>For perceived risk question was restricted to people who did not believe they had already had COVID-19. Total numbers completing this item: n=1186 for keyworkers, and n=901 for health and social care or relevant related support workers, n=1311 for non-keyworkers.
Loneliness and perceived risk were measured on a scale of 1 to 10. Positive mood is a score from 6 to 30.

Further regression analyses were performed to examine whether there were differences in psychological variables between health and social care keyworkers and other keyworkers (unadjusted and adjusted models can be seen in the Supplemental Tables 26-49). After controlling for age, gender, whether the participant lived alone, ethnicity, and clinical risk group status we observed that health and social care keyworkers reported significantly lower levels of stress (B = -0.46, 95% CI: [-0.83, -0.09], *p* = 0.014) and loneliness (B = -0.55, 95% CI: [-0.85, -0.24], *p* <.001), but significantly greater perceived risk of getting COVID-19 (B = 0.75, 95% CI: [0.46, 1.05], *p* < .001) compared to other keyworkers. No statistically significant differences were evident for other psychological variables including depression, anxiety, positive mood, and worry about COVID-19.

### Interactions between keyworker status and gender

Regression models examining an interaction between keyworker status (all keyworkers/non-keyworkers) and gender indicated significant interaction effects for stress (*p* = 0.004), anxiety (*p =* 0.02), and loneliness (*p =* 0.001). These showed that while male respondents, on average, had lower levels of stress, anxiety, and loneliness than female respondents, being a keyworker was associated with increases in stress, anxiety, and loneliness in males (see figure 1). In contrast, female keyworkers had lower levels of stress and loneliness than female non-keyworkers and only slightly elevated anxiety levels.

**Figure 1:**
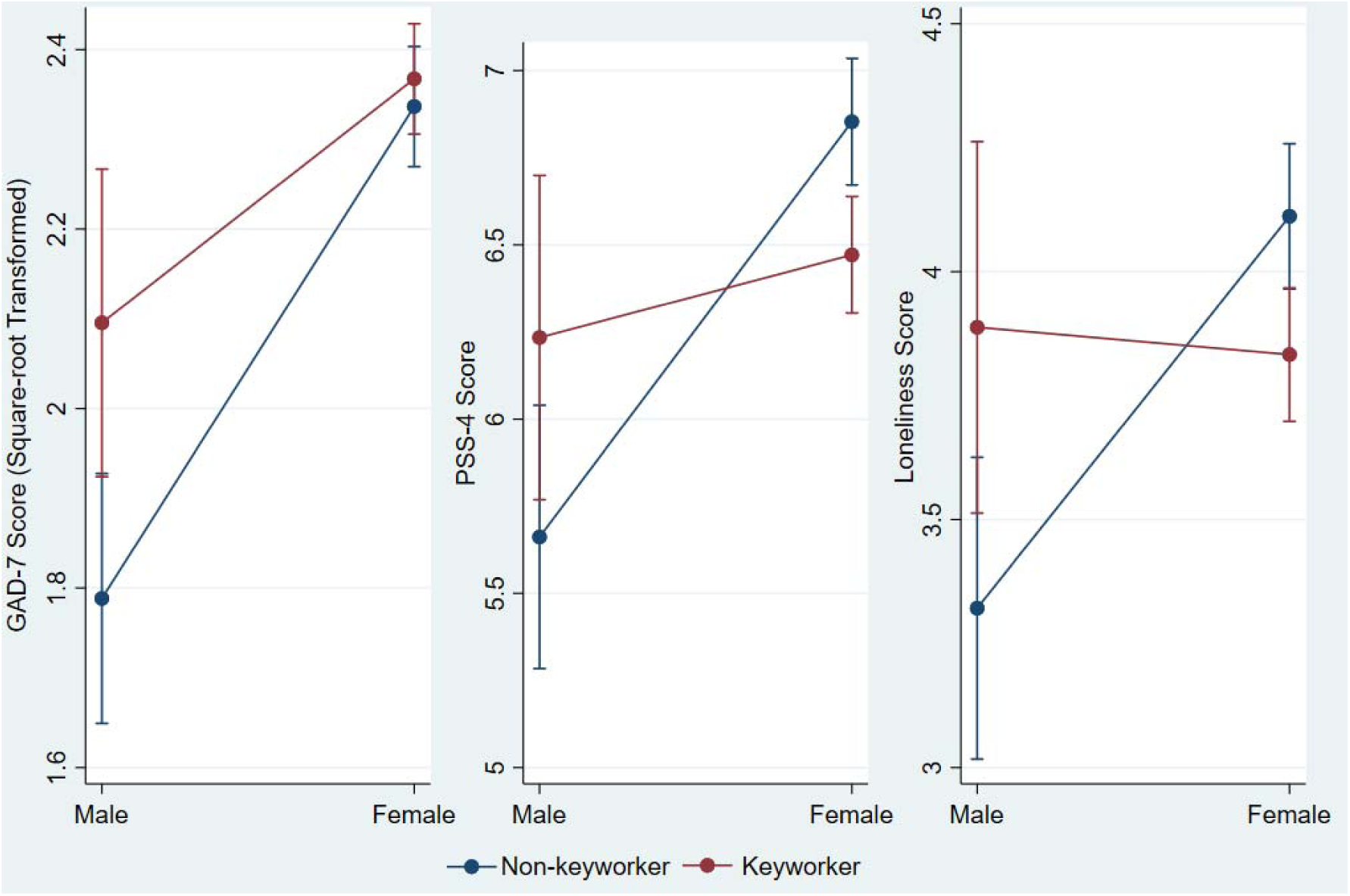
Plots of Interaction between Keyworker Status and Gender for anxiety (GAD-7), stress (PSS-4) and loneliness scores.

### What modifiable and non-modifiable factors are associated with mental health in keyworkers?

In multivariable linear regressions we examined the strength of the association between age, gender (male/female) and ethnicity (white/BAME), whether participants lived alone, clinical risk group status and mental health outcomes within keyworkers (see Table 5). Younger age and being in the “most at risk” group was significantly associated with higher levels of depression, anxiety and stress. Greater anxiety and depression (but not stress) was also observed in female respondents and those in the “increased risk” group. Living alone was associated with significantly higher levels of depression, but not anxiety or stress. There were no significant associations with ethnicity. The amount of variance accounted for by these variables was, however, modest (ranging from 4-6%).

**Table 5:** Multivariable linear regression models showing associations between demographics and depression, anxiety and stress scores for all keyworkers

| | Regression coefficient<br>(B) | 95% CI<br>Lower | 95% CI<br>Upper | $\beta$ | *p* |
| --- | --- | --- | --- | --- | --- |
| **Depression (PHQ-9) Score <sup>a</sup>** |  |  |  |  |  |
| Age (per decade) | -0.22 | -0.27 | -0.17 | -0.22 | <0.001*** |
| Female | 0.22 | 0.04 | 0.40 | 0.06 | 0.02* |
| Live alone | 0.20 | 0.02 | 0.38 | 0.05 | 0.03* |
| BAME background | -0.02 | -0.21 | 0.18 | -0.00 | 0.85 |
| Risk Group <sup>b</sup> |  |  |  |  |  |
| Most at Risk | 0.44 | 0.15 | 0.74 | 0.07 | 0.003** |
| Increased Risk | 0.36 | 0.19 | 0.52 | 0.10 | <0.001*** |
| Observations | 1,556 |  |  |  |  |
| Adjusted R-squared | 0.06 |  |  |  |  |
| **Anxiety (GAD-7) Total Score <sup>a</sup>** |  |  |  |  |  |
| Age (per decade) | -0.22 | -0.27 | -0.17 | -0.22 | <0.001*** |
| Female | 0.27 | 0.08 | 0.45 | 0.07 | 0.004** |
| Live alone | -0.10 | -0.28 | 0.08 | -0.03 | 0.29 |
| BAME background | 0.03 | -0.17 | 0.22 | 0.01 | 0.81 |
| Risk Group <sup>b</sup> |  |  |  |  |  |
| Most at Risk | 0.34 | 0.04 | 0.65 | 0.06 | 0.03* |
| Increased Risk | 0.28 | 0.10 | 0.45 | 0.08 | 0.002** |
| Observations | 1,556 |  |  |  |  |
| Adjusted R-squared | 0.06 |  |  |  |  |
| **Stress (PSS-4) Score** |  |  |  |  |  |
| Age (per decade) | -0.49 | -0.63 | -0.36 | -0.18 | <0.001*** |
| Female | 0.20 | -0.29 | 0.69 | 0.02 | 0.43 |
| Live alone | 0.32 | -0.17 | 0.81 | 0.03 | 0.20 |
| BAME background | 0.14 | -0.40 | 0.67 | 0.01 | 0.61 |
| Risk Group <sup>b</sup> |  |  |  |  |  |
| Most at Risk | 1.01 | 0.20 | 1.82 | 0.06 | 0.01* |
| Increased Risk | 0.42 | -0.04 | 0.88 | 0.05 | 0.07 |
| Observations | 1,556 |  |  |  |  |
| Adjusted R-squared | 0.04 |  |  |  |  |

\*\*\* p<0.001, \*\* p<0.01, \* p<0.05
<sup>a</sup> A square-root transformation was applied to the dependent variable.
<sup>b</sup> Comparison reference group "I am in neither risk category".

### Psychological characteristics associated with mental health outcomes in keyworkers

Further multivariable linear regressions were used to explore whether other psychological characteristics were associated with stress, anxiety and depression among keyworkers. The characteristics considered were: perceived loneliness, positive mood, worry about getting COVID-19, perceived risk of COVID-19, and whether participants were supporting others outside their immediate family.

Table 6 shows that within keyworkers, greater loneliness, greater than occasional worry about COVID-19, and lower positive mood were all significantly associated with increased depression, anxiety and stress scores. Perceived risk of COVID-19 was also significantly positively associated with stress and anxiety scores. Supporting others outside of the immediate family was associated with greater levels of stress. The models including these variables accounted for a much greater proportion of variance in these mental health outcomes (between 49-53%) than models considering demographic characteristics alone.

**Table 6:** Multivariable linear regression models showing associations between potentially modifiable factors and depression scores in all keyworkers

| | Regression coefficient (B) | 95% CI<br>Lower | 95% CI<br>Upper | $\beta$ | p |
| --- | --- | --- | --- | --- | --- |
| **Depression (PHQ-9) Total Score <sup>a</sup>** |  |  |  |  |  |
| Age (per decade) | -0.13 | -0.17 | -0.09 | -0.13 | <0.001*** |
| Female | 0.15 | -0.00 | 0.30 | 0.04 | 0.05 |
| Live Alone | -0.16 | -0.32 | -0.01 | -0.04 | 0.04* |
| BAME background | -0.09 | -0.25 | 0.07 | -0.02 | 0.28 |
| Risk Group <sup>b</sup> |  |  |  |  |  |
| Most at Risk | 0.30 | 0.05 | 0.54 | 0.05 | 0.02* |
| Increased Risk | 0.27 | 0.12 | 0.41 | 0.08 | <0.001*** |
| Perceived loneliness (per unit) | 0.10 | 0.08 | 0.13 | 0.24 | <0.001*** |
| Positive mood (per unit) | -0.12 | -0.13 | -0.11 | -0.49 | <0.001*** |
| COVID-19 worry <sup>c</sup> |  |  |  |  |  |
| No worry | 0.11 | -0.04 | 0.26 | 0.03 | 0.14 |
| Much of time | 0.29 | 0.15 | 0.43 | 0.09 | <0.001*** |
| Most of time | 0.27 | 0.02 | 0.52 | 0.05 | 0.03* |
| Perceived risk of COVID-19 (per unit) | 0.01 | -0.01 | 0.04 | 0.02 | 0.25 |
| Supporting Others (yes/no) | 0.01 | -0.09 | 0.12 | 0.01 | 0.78 |
| Observations | 1,184 |  |  |  |  |
| Adjusted R-squared | 0.52 |  |  |  |  |
| **Anxiety (GAD-7) Total Score <sup>a</sup>** |  |  |  |  |  |
| Age (per decade) | -0.14 | -0.19 | -0.10 | -0.14 | <0.001*** |
| Female | 0.15 | -0.01 | 0.31 | 0.04 | 0.06 |
| Live Alone | -0.29 | -0.45 | -0.13 | -0.08 | <0.001*** |
| BAME background | -0.09 | -0.26 | 0.08 | -0.02 | 0.30 |
| Risk Group <sup>b</sup> |  |  |  |  |  |
| Most at Risk | 0.08 | -0.17 | 0.34 | 0.01 | 0.53 |
| Increased Risk | 0.10 | -0.05 | 0.25 | 0.03 | 0.20 |
| Perceived loneliness (per unit) | 0.05 | 0.02 | 0.07 | 0.10 | <0.001*** |
| Positive mood (per unit) | -0.12 | -0.13 | -0.10 | -0.48 | <0.001*** |
| COVID-19 worry <sup>c</sup> |  |  |  |  |  |
| No worry | -0.02 | -0.17 | 0.14 | -0.00 | 0.83 |
| Much of time | 0.65 | 0.50 | 0.79 | 0.19 | <0.001*** |
| Most of time | 0.91 | 0.65 | 1.17 | 0.15 | <0.001*** |
| Perceived risk of COVID-19 (per unit) | 0.04 | 0.01 | 0.06 | 0.07 | 0.002** |
| Supporting Others (yes/no) | 0.08 | -0.03 | 0.19 | 0.03 | 0.15 |
| Observations | 1,184 |  |  |  |  |
| Adjusted R-squared | 0.49 |  |  |  |  |
| **Stress (PSS-4) Total Score** |  |  |  |  |  |
| Age (per decade) | -0.27 | -0.37 | -0.16 | -0.10 | <0.001*** |
| Female | -0.04 | -0.44 | 0.36 | -0.00 | 0.84 |
| Live Alone | -0.39 | -0.80 | 0.02 | -0.04 | 0.06 |
| BAME background | 0.04 | -0.39 | 0.47 | 0.00 | 0.86 |
| Risk Group <sup>b</sup> |  |  |  |  |  |
| Most at Risk | 0.40 | -0.24 | 1.04 | 0.02 | 0.22 |
| Increased Risk | 0.00 | -0.37 | 0.38 | 0.00 | 0.99 |
| Perceived loneliness (per unit) | 0.17 | 0.11 | 0.22 | 0.14 | <0.001*** |
| Positive mood (per unit) | -0.37 | -0.40 | -0.34 | -0.59 | <0.001*** |
| COVID-19 worry <sup>c</sup> |  |  |  |  |  |
| No worry | 0.21 | -0.18 | 0.59 | 0.02 | 0.29 |
| Much of time | 0.41 | 0.04 | 0.78 | 0.05 | 0.03* |
| Most of time | 0.91 | 0.26 | 1.56 | 0.06 | 0.01** |
| Perceived risk of COVID-19 (per unit) | 0.08 | 0.02 | 0.14 | 0.06 | 0.01** |
| Supporting Others (yes/no) | 0.40 | 0.13 | 0.66 | 0.06 | 0.004** |
| Observations | 1,184 |  |  |  |  |
| Adjusted R-squared | 0.53 |  |  |  |  |

\*\*\* p<0.001, \*\* p<0.01, \* p<0.05
<sup>a</sup> A square-root transformation was applied to the dependent variable.
<sup>b</sup> Comparison reference group "I am in neither risk category".
<sup>c</sup> Comparison reference group "I occasionally worry about getting COVID-19".

## Discussion

In this study we examined the mental health impact of the early stages of the COVID-19 pandemic on keyworkers in the UK. Our findings show that keyworkers’ mental health during the early stages of the pandemic was considerably poorer than normative pre-pandemic levels for the wider population. Mean depression and anxiety scores were more than double those measured in the general population pre-pandemic, and stress levels (at least in males) were also significantly raised. Approximately a third of keyworkers met the NHS threshold for referral to high intensity IAPT support (score ≥ 10) on the grounds of depression, and a quarter of keyworkers on the grounds of anxiety. These findings are consistent with from previous viral outbreaks such as SARS and MERS, which resulted in substantial increases in mental health morbidity among frontline workers (Cabello et al., 2020). Younger keyworkers and those in a clinically increased risk groups had the worst levels of mental health and were significantly more likely to be experiencing moderate or higher levels of depression and anxiety. These findings are congruent with other emerging evidence worldwide of the significant deleterious mental health impact from the COVID-19 pandemic (Jia et al., 2020; Rajkumar, 2020; Torales, O’Higgins, Castaldelli-Maia, & Ventriglio, 2020; W. R. Zhang et al., 2020; Y. Zhang & Ma, 2020) and provide further weight to the calls made already for greater mental health support for keyworkers (Kinman et al., 2020; Rana et al., 2020; Sim, 2020; The Lancet, 2020; Xiang et al., 2020).

While non-keyworkers also experienced comparable levels of poor mental health, we found keyworkers perceived they were at greater risk from COVID-19 and were more likely to frequently worry about COVID-19 than non-keyworkers, a finding consistent with similarly timed survey data from the Office for National Statistics (Office for National Statistics, 2020). These increased perceptions of risk and worry may be explained by the greater salience of COVID-19 risks for keyworkers because of their frontline position. However, contextually, it is also worth reflecting that during this period there was a highly publicised lack of access to personal protective equipment for keyworkers in the UK (World Health Organization, 2020b) and this may also have contributed to increased perceived, and actual, risk and worry about COVID-19.

Yet, despite these greater concerns around COVID-19 specific risks, we found keyworkers did not significantly differ from non-keyworkers in terms of levels of depression, anxiety, positive mood or loneliness. While this may appear counterintuitive, we speculate that, both keyworkers and non-keyworkers are facing substantial challenges that account for similar increases in psychological morbidity, albeit the specifics of these challenges most likely differ. For example, keyworkers may have greater concerns around their personal risk of being exposed to the virus, but conversely they can continue to work, which is known to be protective for mental health (Modini et al., 2016). They may also benefit from having a greater semblance of continuing normality, public appreciation, and feeling they are contributing to the crisis. This latter point is supported by our finding that health and social care keyworkers, who arguably received the most public acknowledgement for their role during this time, were less lonely and stressed than other types of keyworkers – despite perceiving greater risk from COVID-19. Further, healthcare workers have previously been shown to have greater decision latitude and job control than many other kinds of keyworkers, which may also explain lower stress among keyworkers (Karasek, Baker, Marxer, Ahlbom, & Theorell, 1981). For non-keyworkers, perceived risk of COVID-19 may be lower than that of keyworkers, but they are likely dealing with a range of different stressors potentially including food and income security, trying to home-school children, and being cut off from the vast majority of face-to-face social contacts.

It is noteworthy that we observed significant gender by keyworker status interactions, such that for males: being a keyworker was associated with poorer psychological wellbeing compared to male non-keyworkers (in terms of stress, anxiety, and loneliness). Female keyworkers were less stressed, and less lonely than female non-keyworkers, albeit still having higher mean levels of anxiety and stress compared to male counterparts. While a causal examination of these differences goes beyond the scope of the data reported here, one potential explanation for this may be the disproportionate burden of the pandemic on female non-keyworkers, particularly those who are parents. Evidence collected during the pandemic indicates mothers working from home were more frequently interrupted by children during paid working hours than fathers, had greater domestic responsibilities even where they worked more paid hours, and were more likely to have lost, quit, or been furloughed from their jobs than fathers (Andrew et al., 2020). Such disparities, and concomitant negative impacts on mental health, may be less prevalent for male and female keyworkers, at least during working hours.

Within keyworkers, while some demographic factors (in particular being younger and being in a clinically increased risk group) were associated with poorer mental health outcomes, it is noteworthy that regression models including demographic factors accounted for only 4-6% of the variation in depression, anxiety, and stress levels. In contrast, models including potentially modifiable psychological factors, accounted for between 49-53% of this variation. Specifically, greater positive mood (which is known to be associated with greater psychological resilience; Tugade, Fredrickson, & Feldman Barrett, 2004), reduced loneliness, and worrying less about COVID-19 were all associated with better mental health outcomes in keyworkers. Greater perceived risk of getting COVID-19 was associated with higher levels of anxiety and stress, but not depression. While it is not possible at this stage to determine any causal relationship between these factors, these findings could indicate potential for targeting these factors with psychological interventions. Some early interventions have already been rapidly developed for this purpose (e.g., Blake, Bermingham, Johnson, & Tabner, 2020), but wider policy, structural, environmental, and public health interventions may also play an important role in reducing risk as well as the psychological burdens placed on keyworkers. Examples could include: greater provision of required personal protective equipment, altered shift patterns or role rotations, increases to renumeration, or adopting supportive peer and supervision structures such as “buddy systems” as advocated by the World Health Organisation (2020a) guidance for mental wellbeing of healthcare workers during the pandemic.

### Strengths and Limitations

In this study we report on the mental wellbeing of over 1,500 keyworkers during the early phases of the COVID-19 restrictions in the UK. The cohort was set up and recruited rapidly in response to the COVID-19 pandemic. As such, this is one of only a few studies that can contribute to the relatively sparse mental health data available from this time in the UK and will prove useful for understanding the trajectory of the pandemic’s effect on mental health moving forward. However, it is important to acknowledge the study’s limitations. First, the cross-sectional analyses reported here, which represent data collected during the first wave of a longitudinal community cohort study, only provide a snapshot of the keyworkers’ mental health during April 2020. As such, our findings cannot speak to the persistence or otherwise of these elevated levels of distress. This is important to track because while elevated stress and feelings of pressure may be a normal response to the pandemic (World Health Organization, 2020a), longer term distress can be particularly detrimental to both psychological and physical health (J. Cohen, 2000; Juster, McEwen, & Lupien, 2010; Mulligan et al., 2014; O’Connor, Thayer, & Vedhara, 2020). Recent reports have noted levels of depression, anxiety and stress in frontline workers remained high during May 2020 (Couper et al., 2020), and future waves from the present cohort study will further contribute to this knowledge base.

A further limitation of this work concerns sampling bias. Demographically, while for many of the variables measured, this cohort is broadly representative of the UK population (Jia et al., 2020), women were over-represented in common with other online survey studies of mental health (e.g., Owen et al., 2014). Further, given the self-selecting nature of participation, it may be that those experiencing greater distress were more drawn to participate in the study, in turn contributing to our findings of poor mental health in keyworkers. Conversely, those keyworkers experiencing the highest levels of mental health difficulties might reasonably be suspected to have been less likely to participate in the study, due to the already considerable demands placed upon them. These sampling biases are not easily unpicked, and readers should therefore be mindful of these issues when interpreting findings.

### Concluding Remarks

The data presented above demonstrate the substantial early negative mental health impacts of the COVID-19 pandemic on both UK keyworkers and non-keyworkers. Younger and clinically vulnerable keyworkers are most at risk of experiencing poor mental health and may be most able to benefit from interventions that seek to mitigate the negative mental health impacts of the pandemic. Addressing loneliness, worry about COVID-19, and increasing positive mood may be particularly beneficial within keyworkers – in which psychological, environmental, and policy interventions can all potentially play a role.

## Data Availability

Data are available upon reasonable request. Data will be deposited in the University of Nottingham data archive. Access to this dataset will be embargoed for a period of 12 months to permit planned analyses of the dataset. After that it may be shared with the consent of the Chief Investigator. Extra data is available by contacting.

## Notes

### Competing Interest Statement

The authors have declared no competing interest.

### Funding Statement

KA was supported by funding from the National Institute for Health Research School for Primary Care Research (NIHR SPCR). The views expressed are those of the author(s) and not necessarily those of the NIHR, the NHS or the Department of Health. TC acknowledges the financial support of the Department of Health via the National Institute for Health Research (NIHR) Specialist Biomedical Research Centre for Mental Health award to the South London and Maudsley NHS Foundation Trust (SLaM) and the Institute of Psychiatry at King's College London.
CC acknowledges support from the National Institute for Health Research (NIHR) Nottingham Biomedical Research Centre. The views expressed are those of the authors and not necessarily those of the NHS, the NIHR or the Department of Health and Social Care.

### Author Declarations

Ethics and research governance for the study was granted from the University of Nottingham Faculty of Medicine and Health Sciences (ref: 506-2003) and the NHS Health Research Authority (ref: 20/HRA/1858).

