## Supplemental Appendix for "Mental Health of Keyworkers in the UK during the COVID-19 Pandemic: a Cross-sectional Analysis of a Community Cohort"

Supplemental Table 1: Survey item wording

|  | **Question/scale** | **Response(s)** |
| --- | --- | --- |
| Gender* | What was your gender at birth? | Male |
|  |  | Female |
|  |  | Other |
|  |  | Prefer not to say |
| Age | How old are you? | ·· |
| Ethnicity* | What is your ethnicity | White – British, Irish, other |
|  |  | Asian/Asian British – Indian, Pakistani, Bangladeshi, other |
|  |  | Black/Black British – Caribbean, African, other |
|  |  | Chinese/Chinese British |
|  |  | Mixed race – White and Black/Black British |
|  |  | Middle Eastern/Middle Eastern British – Arab, Turkish, other |
|  |  | Mixed race – other |
|  |  | Other ethnic group |
|  |  | Prefer not to say |
| Key-worker status | Are you currently fulfilling any of the government’s identified ‘key worker’ roles? | Health, social care ore relevant related support worker |
|  |  | Teacher or childcare worker still travelling in to work |
|  |  | Transport worker still travelling in to work |
|  |  | Food chain worker (e.g. production, sale, delivery) |
|  |  | Key public services worker (e.g. justice staff, religious staff, public service journalist or mortuary worker) |
|  |  | Local or national government worker delivering essential public services |
|  |  | Utility worker (e.g. energy, sewerage, postal service) |
|  |  | Public safety or national security worker |
|  |  | Worker involved in medicines or protective equipment production or distribution |
|  |  | Other ‘key worker’ role not listed |
|  |  | None of these |
| Living alone/with others | Do you live with someone? | Yes |
|  |  | No |
| Recognised risk group for COVID-19 | Which of these 3 COVID-19 risk groups do you think you are in? | I am most at risk (e.g., suffering from advanced cancer, severe asthma/COPD, etc.) |
|  |  | I am at increased risk (e.g., being pregnant, aged over 70, etc.) |
|  |  | I am in neither risk category. |
| Perceived loneliness^†^ | On a scale of 1-10, how lonely have you felt over the past 2 weeks? | 1 (Not at all lonely) - 10 (Extremely lonely) |
| Perceived risk of COVID-19 | On a scale of 1-10, what do you believe your risk of getting COVID-19 is? | 1 (I don’t think I will get it) - 10 (I know I will most certainly get it) |
| COVID-19 worry | Please read the following statements carefully and then select the one which best describe how you have felt over the past 2 weeks. | I do not worry about getting COVID-19. |
|  |  | I occasionally worry about getting COVID-19. |
|  |  | I spend much of my time worrying about getting COVID-19. |
|  |  | I spend most of my time worrying about getting COVID-19. |

*Gender and ethnicity were treated as binary variables in all analyses: gender (male, female), ethnicity (white British, non-white British).

**Comparing All Keyworkers and Non-Keyworkers**

Depression

Supplemental Table 2: Unadjusted Regression Model Examining Effect of Keyworker Status on PHQ-9 Total Score

|  | **Regression coefficient (B)** | **95% CI Lower** | **95% CI Upper** | ***β*** | ***p*** |
| --- | --- | --- | --- | --- | --- |
| Keyworker | 0.06 | -0.03 | 0.15 | 0.02 | 0.18 |
| Observations | 2,994 |  |  |  |  |
| Adjusted R-squared | 0.00 |  |  |  |  |

*** p<0.001, ** p<0.01, * p<0.05

Supplemental Table 3: Adjusted Regression Model Examining Effect of Keyworker Status on PHQ-9 Total Score

|  | **Regression coefficient (B)** | **95% CI Lower** | **95% CI Upper** | ***β*** | ***p*** |
| --- | --- | --- | --- | --- | --- |
| Age (per decade) | -0.28 | -0.31 | -0.26 | -0.33 | <.001*** |
| Gender | 0.36 | 0.24 | 0.47 | 0.11 | <.001*** |
| Live alone | 0.34 | 0.21 | 0.46 | 0.09 | <.001*** |
| BAME Background | 0.03 | -0.11 | 0.16 | 0.01 | 0.71 |
| Risk Group |  |  |  |  |  |
| Most at Risk | 0.57 | 0.36 | 0.78 | 0.09 | <.001*** |
| Increased Risk | 0.31 | 0.19 | 0.43 | 0.09 | <.001*** |
| Keyworker | 0.06 | -0.02 | 0.14 | 0.02 | 0.16 |
| Observations | 2,990 |  |  |  |  |
| Adjusted R-squared | 0.13 |  |  |  |  |

*** p<0.001, ** p<0.01, * p<0.05

Supplemental Table 4: Adjusted Regression Model Examining Interaction Between Keyworker Status and Gender on PHQ-9 Total Score

|  | **Regression coefficient (B)** | **95% CI Lower** | **95% CI Upper** | ***β*** | ***p*** |
| --- | --- | --- | --- | --- | --- |
| Age (per decade) | -0.28 | -0.31 | -0.25 | -0.33 | <.001*** |
| Gender | 0.45 | 0.30 | 0.60 | 0.13 | <.001*** |
| Live alone | 0.34 | 0.21 | 0.46 | 0.09 | <.001*** |
| BAME Background | 0.03 | -0.11 | 0.16 | 0.01 | 0.71 |
| Risk Group |  |  |  |  |  |
| Most at Risk | 0.57 | 0.36 | 0.78 | 0.09 | <.001*** |
| Increased Risk | 0.31 | 0.19 | 0.42 | 0.09 | <.001*** |
| Keyworker | 0.24 | 0.03 | 0.45 | 0.10 | 0.03* |
| Gender#Keyworker | -0.21 | -0.44 | 0.02 | -0.09 | 0.07 |
| Observations | 2,990 |  |  |  |  |
| Adjusted R-squared | 0.13 |  |  |  |  |

*** p<0.001, ** p<0.01, * p<0.05

Supplemental Table 5: Unadjusted Regression Model Examining Effect of Keyworker Status on PHQ-9 Score ≥ 10

|  | **Odds Ratio** | **95% CI Lower** | **95% CI Upper** | ***β*** | ***p*** |
| --- | --- | --- | --- | --- | --- |
| Keyworker | 1.07 | 0.92 | 1.25 | 0.08 | 0.36 |
| Observations | 2,994 |  |  |  |  |
| Pseudo R-squared | 0.000222 |  |  |  |  |

*** p<0.001, ** p<0.01, * p<0.05

Supplemental Table 6: Adjusted Regression Model Examining Effect of Keyworker Status on PHQ-9 Score ≥ 10

|  | **Odds Ratio** | **95% CI Lower** | **95% CI Upper** | ***β*** | ***p*** |
| --- | --- | --- | --- | --- | --- |
| Age (per decade) | 0.67 | 0.63 | 0.71 | -1.22 | <.001*** |
| Gender | 1.49 | 1.18 | 1.89 | 0.30 | <.001*** |
| Live alone | 1.57 | 1.23 | 1.99 | 0.32 | <.001*** |
| BAME Background | 1.15 | 0.89 | 1.49 | 0.09 | 0.30 |
| Risk Group |  |  |  |  |  |
| Most at Risk | 1.99 | 1.34 | 2.96 | 0.28 | <.001*** |
| Increased Risk | 1.74 | 1.39 | 2.17 | 0.42 | <.001*** |
| Keyworker | 1.13 | 0.96 | 1.32 | 0.13 | 0.15 |
| Observations | 2,990 |  |  |  |  |
| Pseudo R-squared | 0.0599 |  |  |  |  |

*** p<0.001, ** p<0.01, * p<0.05

Supplemental Table 7: Adjusted Regression Model Examining Effect Interaction Between Keyworker Status and Gender on PHQ-9 Score ≥ 10

|  | **Odds Ratio** | **95% CI Lower** | **95% CI Upper** | ***β*** | ***p*** |
| --- | --- | --- | --- | --- | --- |
| Age (per decade) | 0.67 | 0.63 | 0.71 | -1.22 | <.001*** |
| Gender | 1.38 | 1.01 | 1.88 | 0.25 | 0.04* |
| Live alone | 1.57 | 1.23 | 1.99 | 0.32 | <.001*** |
| BAME Background | 1.15 | 0.89 | 1.49 | 0.09 | 0.30 |
| Risk Group |  |  |  |  |  |
| Most at Risk | 2.00 | 1.35 | 2.97 | 0.28 | <.001*** |
| Increased Risk | 1.74 | 1.39 | 2.17 | 0.42 | <.001*** |
| Keyworker | 0.96 | 0.62 | 1.50 | -0.04 | 0.86 |
| Gender#Keyworker | 1.20 | 0.75 | 1.93 | 0.19 | 0.45 |
| Observations | 2,990 |  |  |  |  |
| Pseudo R-squared | 0.0601 |  |  |  |  |

*** p<0.001, ** p<0.01, * p<0.05

Anxiety

Supplemental Table 8: Unadjusted Regression Model Examining Effect of Keyworker Status on GAD-7 Total Score

|  | **Regression coefficient (B)** | **95% CI Lower** | **95% CI Upper** | ***β*** | ***p*** |
| --- | --- | --- | --- | --- | --- |
| Keyworker | 0.09 | -0.00 | 0.17 | 0.04 | 0.05 |
| Observations | 2,994 |  |  |  |  |
| Adjusted R-squared | 0.00 |  |  |  |  |

*** p<0.001, ** p<0.01, * p<0.05

Supplemental Table 9: Adjusted Regression Model Examining Effect of Keyworker Status on GAD-7 Total Score

|  | **Regression coefficient (B)** | **95% CI Lower** | **95% CI Upper** | ***β*** | ***p*** |
| --- | --- | --- | --- | --- | --- |
| Age (per decade) | -0.24 | -0.27 | -0.21 | -0.28 | <.001*** |
| Gender | 0.43 | 0.31 | 0.55 | 0.13 | <.001*** |
| Live alone | -0.00 | -0.13 | 0.13 | -0.00 | 0.97 |
| BAME Background | 0.02 | -0.13 | 0.16 | 0.00 | 0.82 |
| Risk Group |  |  |  |  |  |
| Most at Risk | 0.44 | 0.22 | 0.66 | 0.07 | <.001*** |
| Increased Risk | 0.27 | 0.15 | 0.39 | 0.08 | <.001*** |
| Keyworker | 0.07 | -0.01 | 0.16 | 0.03 | 0.10 |
| Observations | 2,990 |  |  |  |  |
| Adjusted R-squared | 0.10 |  |  |  |  |

*** p<0.001, ** p<0.01, * p<0.05

Supplemental Table 10: Adjusted Regression Model Examining Interaction Between Keyworker Status and Gender on GAD-7 Total Score

|  | **Regression coefficient (B)** | **95% CI Lower** | **95% CI Upper** | ***β*** | ***p*** |
| --- | --- | --- | --- | --- | --- |
| Age (per decade) | -0.24 | -0.27 | -0.21 | -0.28 | <.001*** |
| Gender | 0.55 | 0.39 | 0.70 | 0.16 | <.001*** |
| Live alone | -0.00 | -0.13 | 0.12 | -0.00 | 0.94 |
| BAME Background | 0.02 | -0.13 | 0.16 | 0.00 | 0.81 |
| Risk Group |  |  |  |  |  |
| Most at Risk | 0.43 | 0.21 | 0.65 | 0.07 | <.001*** |
| Increased Risk | 0.26 | 0.14 | 0.38 | 0.08 | <.001*** |
| Keyworker | 0.31 | 0.09 | 0.53 | 0.13 | 0.01** |
| Gender#Keyworker | -0.28 | -0.51 | -0.04 | -0.11 | 0.02* |
| Observations | 2,990 |  |  |  |  |
| Adjusted R-squared | 0.10 |  |  |  |  |

*** p<0.001, ** p<0.01, * p<0.05

Supplemental Table 11: Unadjusted Regression Model Examining Effect of Keyworker Status on GAD-7 Score ≥ 10

|  | **Odds Ratio** | **95% CI Lower** | **95% CI Upper** | ***β*** | ***p*** |
| --- | --- | --- | --- | --- | --- |
| Keyworker | 1.08 | 0.92 | 1.27 | 0.09 | 0.36 |
| Observations | 2,994 |  |  |  |  |
| Pseudo R-squared | 0.000246 |  |  |  |  |

*** p<0.001, ** p<0.01, * p<0.05

Supplemental Table 12: Adjusted Regression Model Examining Effect of Keyworker Status on GAD-7 Score ≥ 10

|  | **Odds Ratio** | **95% CI Lower** | **95% CI Upper** | ***β*** | ***p*** |
| --- | --- | --- | --- | --- | --- |
| Age (per decade) | 0.70 | 0.66 | 0.75 | -1.13 | <.001*** |
| Gender | 1.64 | 1.27 | 2.12 | 0.40 | <.001*** |
| Live alone | 1.04 | 0.80 | 1.36 | 0.03 | 0.75 |
| BAME Background | 1.15 | 0.88 | 1.51 | 0.10 | 0.30 |
| Risk Group |  |  |  |  |  |
| Most at Risk | 1.79 | 1.19 | 2.68 | 0.25 | 0.01** |
| Increased Risk | 1.38 | 1.09 | 1.74 | 0.25 | 0.01** |
| Keyworker | 1.11 | 0.94 | 1.31 | 0.12 | 0.22 |
| Observations | 2,990 |  |  |  |  |
| Pseudo R-squared | 0.0456 |  |  |  |  |

*** p<0.001, ** p<0.01, * p<0.05

Supplemental Table 13: Adjusted Regression Model Examining Interaction Between Keyworker Status and Gender on GAD-7 Score ≥ 10

|  | **Odds Ratio** | **95% CI Lower** | **95% CI Upper** | ***β*** | ***p*** |
| --- | --- | --- | --- | --- | --- |
| Age (per decade) | 0.70 | 0.66 | 0.75 | -1.13 | <.001*** |
| Gender | 2.01 | 1.42 | 2.86 | 0.56 | <.001*** |
| Live alone | 1.04 | 0.80 | 1.35 | 0.03 | 0.76 |
| BAME Background | 1.16 | 0.88 | 1.51 | 0.10 | 0.29 |
| Risk Group |  |  |  |  |  |
| Most at Risk | 1.77 | 1.18 | 2.66 | 0.25 | 0.01** |
| Increased Risk | 1.37 | 1.08 | 1.73 | 0.25 | 0.01** |
| Keyworker | 1.65 | 1.02 | 2.65 | 0.56 | 0.04* |
| Gender#Keyworker | 0.64 | 0.38 | 1.06 | -0.51 | 0.08 |
| Observations | 2,990 |  |  |  |  |
| Pseudo R-squared | 0.0465 |  |  |  |  |

*** p<0.001, ** p<0.01, * p<0.05

Stress

Supplemental Table 14: Unadjusted Regression Model Examining Effect of Keyworker Status on PSS-4 Total Score

|  | **Regression coefficient (B)** | **95% CI Lower** | **95% CI Upper** | ***β*** | ***p*** |
| --- | --- | --- | --- | --- | --- |
| Keyworker | -0.23 | -0.47 | 0.00 | -0.04 | 0.05 |
| Observations | 2,994 |  |  |  |  |
| Adjusted R-squared | 0.00 |  |  |  |  |

*** p<0.001, ** p<0.01, * p<0.05

Supplemental Table 15: Adjusted Regression Model Examining Effect of Keyworker Status on PSS-4 Total Score

|  | **Regression coefficient (B)** | **95% CI Lower** | **95% CI Upper** | ***β*** | ***p*** |
| --- | --- | --- | --- | --- | --- |
| Age (per decade) | -0.55 | -0.63 | -0.46 | -0.23 | <.001*** |
| Gender | 0.79 | 0.47 | 1.11 | 0.09 | <.001*** |
| Live alone | 0.52 | 0.18 | 0.87 | 0.05 | <.01** |
| BAME Background | 0.43 | 0.05 | 0.82 | 0.04 | 0.03* |
| Risk Group |  |  |  |  |  |
| Most at Risk | 1.16 | 0.57 | 1.76 | 0.07 | <.001*** |
| Increased Risk | 0.44 | 0.12 | 0.77 | 0.05 | 0.01** |
| Keyworker | -0.24 | -0.47 | -0.01 | -0.04 | 0.04* |
| Observations | 2,990 |  |  |  |  |
| Adjusted R-squared | 0.07 |  |  |  |  |

*** p<0.001, ** p<0.01, * p<0.05

Supplemental Table 16: Adjusted Regression Model Examining Interaction Between Keyworker Status and Gender on PSS-4 Total Score

|  | **Regression coefficient (B)** | **95% CI Lower** | **95% CI Upper** | ***β*** | ***p*** |
| --- | --- | --- | --- | --- | --- |
| Age (per decade) | -0.55 | -0.63 | -0.46 | -0.23 | <.001*** |
| Gender | 1.19 | 0.77 | 1.61 | 0.13 | <.001*** |
| Live alone | 0.52 | 0.17 | 0.87 | 0.05 | <.01** |
| BAME Background | 0.44 | 0.05 | 0.82 | 0.04 | 0.03* |
| Risk Group |  |  |  |  |  |
| Most at Risk | 1.14 | 0.55 | 1.74 | 0.07 | <.001*** |
| Increased Risk | 0.43 | 0.11 | 0.76 | 0.05 | 0.01** |
| Keyworker | 0.57 | -0.03 | 1.17 | 0.09 | 0.06 |
| Gender#Keyworker | -0.95 | -1.60 | -0.31 | -0.15 | 0.00** |
| Observations | 2,990 |  |  |  |  |
| Adjusted R-squared | 0.07 |  |  |  |  |

*** p<0.001, ** p<0.01, * p<0.05

Loneliness

Supplemental Table 17: Unadjusted Regression Model Examining Effect of Keyworker Status on Loneliness

|  | **Regression coefficient (B)** | **95% CI Lower** | **95% CI Upper** | ***β*** | ***p*** |
| --- | --- | --- | --- | --- | --- |
| Keyworker | -0.19 | -0.39 | 0.01 | -0.03 | 0.06 |
| Observations | 2,994 |  |  |  |  |
| Adjusted R-squared | 0.00 |  |  |  |  |

*** p<0.001, ** p<0.01, * p<0.05

Supplemental Table 18: Adjusted Regression Model Examining Effect of Keyworker Status on Loneliness

|  | **Regression coefficient (B)** | **95% CI Lower** | **95% CI Upper** | ***β*** | ***p*** |
| --- | --- | --- | --- | --- | --- |
| Age (per decade) | -0.53 | -0.60 | -0.46 | -0.27 | <.001*** |
| Gender | 0.44 | 0.18 | 0.70 | 0.06 | <.001*** |
| Live alone | 2.34 | 2.06 | 2.62 | 0.28 | <.001*** |
| BAME Background | 0.26 | -0.05 | 0.58 | 0.03 | 0.10 |
| Risk Group |  |  |  |  |  |
| Most at Risk | 1.21 | 0.73 | 1.69 | 0.08 | <.001*** |
| Increased Risk | 0.41 | 0.15 | 0.67 | 0.05 | <.01** |
| Keyworker | -0.16 | -0.34 | 0.03 | -0.03 | 0.10 |
| Observations | 2,990 |  |  |  |  |
| Adjusted R-squared | 0.14 |  |  |  |  |

*** p<0.001, ** p<0.01, * p<0.05

Supplemental Table 19: Adjusted Regression Model Examining Interaction Between Keyworker Status and Gender on Loneliness

|  | **Regression coefficient (B)** | **95% CI Lower** | **95% CI Upper** | ***β*** | ***p*** |
| --- | --- | --- | --- | --- | --- |
| Age (per decade) | -0.53 | -0.59 | -0.46 | -0.27 | <.001*** |
| Gender | 0.79 | 0.45 | 1.13 | 0.10 | <.001*** |
| Live alone | 2.33 | 2.05 | 2.61 | 0.28 | <.001*** |
| BAME Background | 0.26 | -0.05 | 0.58 | 0.03 | 0.10 |
| Risk Group |  |  |  |  |  |
| Most at Risk | 1.19 | 0.71 | 1.67 | 0.08 | <.001*** |
| Increased Risk | 0.40 | 0.14 | 0.66 | 0.05 | <.01** |
| Keyworker | 0.57 | 0.08 | 1.05 | 0.10 | 0.02* |
| Gender#Keyworker | -0.85 | -1.37 | -0.32 | -0.15 | 0.00** |
| Observations | 2,990 |  |  |  |  |
| Adjusted R-squared | 0.15 |  |  |  |  |

*** p<0.001, ** p<0.01, * p<0.05

Positive Mood

Supplemental Table 20: Unadjusted Regression Model Examining Effect of Keyworker Status on SPANE-PA Score

|  | **Regression coefficient (B)** | **95% CI Lower** | **95% CI Upper** | ***β*** | ***p*** |
| --- | --- | --- | --- | --- | --- |
| Keyworker | 0.04 | -0.32 | 0.41 | 0.00 | 0.81 |
| Observations | 2,994 |  |  |  |  |
| Adjusted R-squared | -0.00 |  |  |  |  |

*** p<0.001, ** p<0.01, * p<0.05

Supplemental Table 21: Adjusted Regression Model Examining Effect of Keyworker Status on SPANE-PA Score

|  | **Regression coefficient (B)** | **95% CI Lower** | **95% CI Upper** | ***β*** | ***p*** |
| --- | --- | --- | --- | --- | --- |
| Age (per decade) | 0.45 | 0.32 | 0.58 | 0.13 | <.001*** |
| Gender | -0.95 | -1.46 | -0.44 | -0.07 | <.001*** |
| Live alone | -1.11 | -1.66 | -0.56 | -0.07 | <.001*** |
| BAME Background | -0.37 | -0.98 | 0.24 | -0.02 | 0.24 |
| Risk Group |  |  |  |  |  |
| Most at Risk | -1.80 | -2.73 | -0.87 | -0.07 | <.001*** |
| Increased Risk | -0.74 | -1.26 | -0.23 | -0.05 | <.01** |
| Keyworker | 0.05 | -0.31 | 0.41 | 0.00 | 0.80 |
| Observations | 2,990 |  |  |  |  |
| Adjusted R-squared | 0.03 |  |  |  |  |

*** p<0.001, ** p<0.01, * p<0.05

Supplemental Table 22: Adjusted Regression Model Examining Interaction Between Keyworker Status and Gender on SPANE-PA Score

|  | **Regression coefficient (B)** | **95% CI Lower** | **95% CI Upper** | ***β*** | ***p*** |
| --- | --- | --- | --- | --- | --- |
| Age (per decade) | 0.45 | 0.32 | 0.58 | 0.13 | <.001*** |
| Gender | -1.15 | -1.81 | -0.48 | -0.08 | <.001*** |
| Live alone | -1.11 | -1.66 | -0.56 | -0.07 | <.001*** |
| BAME Background | -0.37 | -0.98 | 0.24 | -0.02 | 0.24 |
| Risk Group |  |  |  |  |  |
| Most at Risk | -1.79 | -2.72 | -0.86 | -0.07 | <.001*** |
| Increased Risk | -0.74 | -1.25 | -0.23 | -0.05 | <.01** |
| Keyworker | -0.36 | -1.30 | 0.59 | -0.04 | 0.46 |
| Gender#Keyworker | 0.47 | -0.55 | 1.49 | 0.05 | 0.37 |
| Observations | 2,990 |  |  |  |  |
| Adjusted R-squared | 0.03 |  |  |  |  |

*** p<0.001, ** p<0.01, * p<0.05

Perceived Risk from COVID-19

Supplemental Table 23: Unadjusted Regression Model Examining Effect of Keyworker Status on Perceived Risk from COVID-19

|  | **Regression coefficient (B)** | **95% CI Lower** | **95% CI Upper** | ***β*** | ***p*** |
| --- | --- | --- | --- | --- | --- |
| Keyworker | 1.48 | 1.32 | 1.65 | 0.33 | <.001*** |
| Observations | 2,400 |  |  |  |  |
| Adjusted R-squared | 0.11 |  |  |  |  |

*** p<0.001, ** p<0.01, * p<0.05

Supplemental Table 24: Adjusted Regression Model Examining Effect of Keyworker Status on Perceived Risk from COVID-19

|  | **Regression coefficient (B)** | **95% CI Lower** | **95% CI Upper** | ***β*** | ***p*** |
| --- | --- | --- | --- | --- | --- |
| Age (per decade) | 0.01 | -0.05 | 0.07 | 0.01 | 0.69 |
| Gender | 0.07 | -0.17 | 0.31 | 0.01 | 0.56 |
| Live alone | -0.14 | -0.39 | 0.12 | -0.02 | 0.30 |
| BAME Background | 0.00 | -0.28 | 0.29 | 0.00 | 0.98 |
| Risk Group |  |  |  |  |  |
| Most at Risk | 0.49 | 0.06 | 0.92 | 0.04 | 0.03* |
| Increased Risk | 0.28 | 0.04 | 0.53 | 0.05 | 0.02* |
| Keyworker | 1.48 | 1.32 | 1.65 | 0.33 | <.001*** |
| Observations | 2,399 |  |  |  |  |
| Adjusted R-squared | 0.11 |  |  |  |  |

*** p<0.001, ** p<0.01, * p<0.05

Supplemental Table 25: Adjusted Regression Model Examining Interaction Between Keyworker Status and Gender on Perceived Risk from COVID-19

|  | **Regression coefficient (B)** | **95% CI Lower** | **95% CI Upper** | ***β*** | ***p*** |
| --- | --- | --- | --- | --- | --- |
| Age (per decade) | 0.01 | -0.05 | 0.07 | 0.01 | 0.68 |
| Gender | 0.13 | -0.17 | 0.42 | 0.02 | 0.41 |
| Live alone | -0.14 | -0.39 | 0.12 | -0.02 | 0.30 |
| BAME Background | 0.00 | -0.28 | 0.29 | 0.00 | 0.99 |
| Risk Group |  |  |  |  |  |
| Most at Risk | 0.48 | 0.05 | 0.92 | 0.04 | 0.03* |
| Increased Risk | 0.28 | 0.04 | 0.52 | 0.04 | 0.02* |
| Keyworker | 1.61 | 1.16 | 2.06 | 0.36 | 0.00*** |
| Gender#Keyworker | -0.15 | -0.64 | 0.34 | -0.03 | 0.55 |
| Observations | 2,399 |  |  |  |  |
| Adjusted R-squared | 0.11 |  |  |  |  |

*** p<0.001, ** p<0.01, * p<0.05

**Comparing Health and Social Care Keyworkers and Other Keyworkers**

Depression

Supplemental Table 26: Unadjusted Regression Model Examining Effect of Keyworker Status on PHQ-9 Total Score

|  | **Regression coefficient (B)** | **95% CI Lower** | **95% CI Upper** | ***β*** | ***p*** |
| --- | --- | --- | --- | --- | --- |
| Health/Social Keyworker | -0.09 | -0.22 | 0.05 | -0.03 | 0.23 |
| Observations | 1,558 |  |  |  |  |
| Adjusted R-squared | 0.00 |  |  |  |  |

*** p<0.001, ** p<0.01, * p<0.05

Supplemental Table 27: Adjusted Regression Model Examining Effect of Keyworker Status on PHQ-9 Total Score

|  | **Regression coefficient (B)** | **95% CI Lower** | **95% CI Upper** | ***β*** | ***p*** |
| --- | --- | --- | --- | --- | --- |
| Age (per decade) | -0.22 | -0.27 | -0.17 | -0.22 | <.001*** |
| Gender | 0.23 | 0.05 | 0.41 | 0.06 | 0.01* |
| Live alone | 0.20 | 0.02 | 0.38 | 0.06 | 0.03* |
| BAME Background | -0.01 | -0.21 | 0.18 | -0.00 | 0.90 |
| Risk Group |  |  |  |  |  |
| Most at Risk | 0.44 | 0.14 | 0.74 | 0.07 | <.01** |
| Increased Risk | 0.36 | 0.19 | 0.53 | 0.10 | <.001*** |
| Health/Social Keyworker | -0.08 | -0.21 | 0.06 | -0.03 | 0.27 |
| Observations | 1,556 |  |  |  |  |
| Adjusted R-squared | 0.06 |  |  |  |  |

*** p<0.001, ** p<0.01, * p<0.05

Supplemental Table 28: Adjusted Regression Model Examining Interaction Between Keyworker Status and Gender on PHQ-9 Total Score

|  | **Regression coefficient (B)** | **95% CI Lower** | **95% CI Upper** | ***β*** | ***p*** |
| --- | --- | --- | --- | --- | --- |
| Age (per decade) | -0.22 | -0.27 | -0.17 | -0.22 | <.001*** |
| Gender | 0.32 | -0.01 | 0.65 | 0.09 | 0.06 |
| Live alone | 0.20 | 0.02 | 0.38 | 0.05 | 0.03* |
| BAME Background | -0.02 | -0.21 | 0.18 | -0.00 | 0.87 |
| Risk Group |  |  |  |  |  |
| Most at Risk | 0.44 | 0.14 | 0.73 | 0.07 | <.01** |
| Increased Risk | 0.36 | 0.19 | 0.52 | 0.10 | <.001*** |
| Health/Social Keyworker | -0.09 | -0.24 | 0.05 | -0.03 | 0.21 |
| Gender#Health/Social Keyworker | 0.13 | -0.26 | 0.52 | 0.03 | 0.51 |
| Observations | 1,556 |  |  |  |  |
| Adjusted R-squared | 0.06 |  |  |  |  |

*** p<0.001, ** p<0.01, * p<0.05

Supplemental Table 29: Unadjusted Regression Model Examining Effect of Keyworker Status on PHQ-9 Score ≥ 10

|  | **Odds Ratio** | **95% CI Lower** | **95% CI Upper** | ***β*** | ***p*** |
| --- | --- | --- | --- | --- | --- |
| Health/Social Keyworker | 0.81 | 0.63 | 1.04 | -0.19 | 0.09 |
| Observations | 1,558 |  |  |  |  |
| Pseudo R-squared | 0.00142 |  |  |  |  |

*** p<0.001, ** p<0.01, * p<0.05

Supplemental Table 30: Adjusted Regression Model Examining Effect of Keyworker Status on PHQ-9 Score ≥ 10

|  | **Odds Ratio** | **95% CI Lower** | **95% CI Upper** | ***β*** | ***p*** |
| --- | --- | --- | --- | --- | --- |
| Age (per decade) | 0.76 | 0.69 | 0.83 | -0.70 | <.001*** |
| Gender | 1.63 | 1.13 | 2.34 | 0.33 | 0.01** |
| Live alone | 1.40 | 1.00 | 1.95 | 0.23 | 0.05* |
| BAME Background | 1.10 | 0.77 | 1.58 | 0.06 | 0.60 |
| Risk Group |  |  |  |  |  |
| Most at Risk | 1.57 | 0.91 | 2.69 | 0.18 | 0.11 |
| Increased Risk | 1.89 | 1.40 | 2.56 | 0.46 | <.001*** |
| Health/Social Keyworker | 0.80 | 0.62 | 1.03 | -0.20 | 0.08 |
| Observations | 1,556 |  |  |  |  |
| Pseudo R-squared | 0.0324 |  |  |  |  |

*** p<0.001, ** p<0.01, * p<0.05

Supplemental Table 31: Adjusted Regression Model Examining Effect Interaction Between Keyworker Status and Gender on PHQ-9 Score ≥ 10

|  | **Odds Ratio** | **95% CI Lower** | **95% CI Upper** | ***β*** | ***p*** |
| --- | --- | --- | --- | --- | --- |
| Age (per decade) | 0.76 | 0.69 | 0.83 | -0.70 | <.001*** |
| Gender | 1.49 | 0.80 | 2.81 | 0.27 | 0.21 |
| Live alone | 1.40 | 1.01 | 1.95 | 0.23 | 0.05* |
| BAME Background | 1.11 | 0.77 | 1.59 | 0.06 | 0.59 |
| Risk Group |  |  |  |  |  |
| Most at Risk | 1.57 | 0.91 | 2.70 | 0.18 | 0.10 |
| Increased Risk | 1.89 | 1.40 | 2.56 | 0.46 | <.001*** |
| Health/Social Keyworker | 0.81 | 0.62 | 1.06 | -0.19 | 0.12 |
| Gender#Health/Social Keyworker | 0.88 | 0.41 | 1.90 | -0.07 | 0.75 |
| Observations | 1,556 |  |  |  |  |
| Pseudo R-squared | 0.0324 |  |  |  |  |

*** p<0.001, ** p<0.01, * p<0.05

Anxiety

Supplemental Table 32: Unadjusted Regression Model Examining Effect of Keyworker Status on GAD-7 Total Score

|  | **Regression coefficient (B)** | **95% CI Lower** | **95% CI Upper** | ***β*** | ***p*** |
| --- | --- | --- | --- | --- | --- |
| Health/Social Keyworker | -0.07 | -0.21 | 0.07 | -0.02 | 0.33 |
| Observations | 1,558 |  |  |  |  |
| Adjusted R-squared | -0.00 |  |  |  |  |

*** p<0.001, ** p<0.01, * p<0.05

Supplemental Table 33: Adjusted Regression Model Examining Effect of Keyworker Status on GAD-7 Total Score

|  | **Regression coefficient (B)** | **95% CI Lower** | **95% CI Upper** | ***β*** | ***p*** |
| --- | --- | --- | --- | --- | --- |
| Age (per decade) | -0.22 | -0.27 | -0.17 | -0.22 | <.001*** |
| Gender | 0.27 | 0.09 | 0.46 | 0.07 | <.01** |
| Live alone | -0.10 | -0.28 | 0.09 | -0.03 | 0.30 |
| BAME Background | 0.03 | -0.17 | 0.23 | 0.01 | 0.77 |
| Risk Group |  |  |  |  |  |
| Most at Risk | 0.34 | 0.04 | 0.64 | 0.05 | 0.03* |
| Increased Risk | 0.28 | 0.11 | 0.45 | 0.08 | <.01** |
| Health/Social Keyworker | -0.06 | -0.20 | 0.08 | -0.02 | 0.42 |
| Observations | 1,556 |  |  |  |  |
| Adjusted R-squared | 0.06 |  |  |  |  |

*** p<0.001, ** p<0.01, * p<0.05

Supplemental Table 34: Adjusted Regression Model Examining Interaction Between Keyworker Status and Gender on GAD-7 Total Score

|  | **Regression coefficient (B)** | **95% CI Lower** | **95% CI Upper** | ***β*** | ***p*** |
| --- | --- | --- | --- | --- | --- |
| Age (per decade) | -0.22 | -0.27 | -0.17 | -0.22 | <.001*** |
| Gender | 0.39 | 0.06 | 0.73 | 0.10 | 0.02* |
| Live alone | -0.10 | -0.28 | 0.08 | -0.03 | 0.29 |
| BAME Background | 0.03 | -0.17 | 0.23 | 0.01 | 0.80 |
| Risk Group |  |  |  |  |  |
| Most at Risk | 0.34 | 0.03 | 0.64 | 0.05 | 0.03* |
| Increased Risk | 0.28 | 0.10 | 0.45 | 0.08 | <.01** |
| Health/Social Keyworker | -0.08 | -0.23 | 0.07 | -0.03 | 0.29 |
| Gender#Health/Social Keyworker | 0.17 | -0.23 | 0.57 | 0.04 | 0.40 |
| Observations | 1,556 |  |  |  |  |
| Adjusted R-squared | 0.06 |  |  |  |  |

*** p<0.001, ** p<0.01, * p<0.05

Supplemental Table 35: Unadjusted Regression Model Examining Effect of Keyworker Status on GAD-7 Score ≥ 10

|  | **Odds Ratio** | **95% CI Lower** | **95% CI Upper** | ***β*** | ***p*** |
| --- | --- | --- | --- | --- | --- |
| Health/Social Keyworker | 0.83 | 0.64 | 1.07 | -0.18 | 0.15 |
| Observations | 1,558 |  |  |  |  |
| Pseudo R-squared | 0.00114 |  |  |  |  |

*** p<0.001, ** p<0.01, * p<0.05

Supplemental Table 36: Adjusted Regression Model Examining Effect of Keyworker Status on GAD-7 Score ≥ 10

|  | **Odds Ratio** | **95% CI Lower** | **95% CI Upper** | ***β*** | ***p*** |
| --- | --- | --- | --- | --- | --- |
| Age (per decade) | 0.75 | 0.68 | 0.82 | -0.77 | <.001*** |
| Gender | 1.29 | 0.89 | 1.86 | 0.18 | 0.18 |
| Live alone | 0.79 | 0.54 | 1.15 | -0.17 | 0.22 |
| BAME Background | 1.24 | 0.86 | 1.80 | 0.15 | 0.25 |
| Risk Group |  |  |  |  |  |
| Most at Risk | 1.64 | 0.93 | 2.87 | 0.21 | 0.09 |
| Increased Risk | 1.42 | 1.03 | 1.96 | 0.27 | 0.03* |
| Health/Social Keyworker | 0.84 | 0.64 | 1.09 | -0.17 | 0.19 |
| Observations | 1,556 |  |  |  |  |
| Pseudo R-squared | 0.0279 |  |  |  |  |

*** p<0.001, ** p<0.01, * p<0.05

Supplemental Table 37: Adjusted Regression Model Examining Interaction Between Keyworker Status and Gender on GAD-7 Score ≥ 10

|  | **Odds Ratio** | **95% CI Lower** | **95% CI Upper** | ***β*** | ***p*** |
| --- | --- | --- | --- | --- | --- |
| Age (per decade) | 0.75 | 0.68 | 0.82 | -0.78 | <.001*** |
| Gender | 1.68 | 0.85 | 3.33 | 0.37 | 0.13 |
| Live alone | 0.79 | 0.54 | 1.15 | -0.17 | 0.21 |
| BAME Background | 1.23 | 0.85 | 1.79 | 0.14 | 0.27 |
| Risk Group |  |  |  |  |  |
| Most at Risk | 1.63 | 0.93 | 2.85 | 0.21 | 0.09 |
| Increased Risk | 1.42 | 1.03 | 1.95 | 0.27 | 0.03* |
| Health/Social Keyworker | 0.80 | 0.60 | 1.06 | -0.21 | 0.12 |
| Gender#Health/Social Keyworker | 1.48 | 0.66 | 3.32 | 0.24 | 0.34 |
| Observations | 1,556 |  |  |  |  |
| Pseudo R-squared | 0.0284 |  |  |  |  |

*** p<0.001, ** p<0.01, * p<0.05

Stress

Supplemental Table 38: Unadjusted Regression Model Examining Effect of Keyworker Status on PSS-4 Total Score

|  | **Regression coefficient (B)** | **95% CI Lower** | **95% CI Upper** | ***β*** | ***p*** |
| --- | --- | --- | --- | --- | --- |
| Health/Social Keyworker | -0.51 | -0.88 | -0.14 | -0.07 | 0.01** |
| Observations | 1,558 |  |  |  |  |
| Adjusted R-squared | 0.00 |  |  |  |  |

*** p<0.001, ** p<0.01, * p<0.05

Supplemental Table 39: Adjusted Regression Model Examining Effect of Keyworker Status on PSS-4 Total Score

|  | **Regression coefficient (B)** | **95% CI Lower** | **95% CI Upper** | ***β*** | ***p*** |
| --- | --- | --- | --- | --- | --- |
| Age (per decade) | -0.49 | -0.62 | -0.35 | -0.18 | <.001*** |
| Gender | 0.24 | -0.25 | 0.73 | 0.02 | 0.34 |
| Live alone | 0.34 | -0.15 | 0.82 | 0.03 | 0.18 |
| BAME Background | 0.17 | -0.36 | 0.71 | 0.02 | 0.52 |
| Risk Group |  |  |  |  |  |
| Most at Risk | 0.97 | 0.16 | 1.79 | 0.06 | 0.02* |
| Increased Risk | 0.43 | -0.03 | 0.89 | 0.05 | 0.06 |
| Health/Social Keyworker | -0.46 | -0.83 | -0.09 | -0.06 | 0.01* |
| Observations | 1,556 |  |  |  |  |
| Adjusted R-squared | 0.04 |  |  |  |  |

*** p<0.001, ** p<0.01, * p<0.05

Supplemental Table 40: Adjusted Regression Model Examining Interaction Between Keyworker Status and Gender on PSS-4 Total Score

|  | **Regression coefficient (B)** | **95% CI Lower** | **95% CI Upper** | ***β*** | ***p*** |
| --- | --- | --- | --- | --- | --- |
| Age (per decade) | -0.49 | -0.62 | -0.35 | -0.18 | <.001*** |
| Gender | 0.30 | -0.60 | 1.19 | 0.03 | 0.52 |
| Live alone | 0.34 | -0.15 | 0.82 | 0.03 | 0.18 |
| BAME Background | 0.17 | -0.36 | 0.70 | 0.02 | 0.53 |
| Risk Group |  |  |  |  |  |
| Most at Risk | 0.97 | 0.16 | 1.78 | 0.06 | 0.02* |
| Increased Risk | 0.43 | -0.03 | 0.89 | 0.05 | 0.07 |
| Health/Social Keyworker | -0.47 | -0.87 | -0.08 | -0.06 | 0.02* |
| Gender#Health/Social Keyworker | 0.08 | -0.99 | 1.15 | 0.01 | 0.89 |
| Observations | 1,556 |  |  |  |  |
| Adjusted R-squared | 0.04 |  |  |  |  |

*** p<0.001, ** p<0.01, * p<0.05

Loneliness

Supplemental Table 41: Unadjusted Regression Model Examining Effect of Keyworker Status on Loneliness

|  | **Regression coefficient (B)** | **95% CI Lower** | **95% CI Upper** | ***β*** | ***p*** |
| --- | --- | --- | --- | --- | --- |
| Health/Social Keyworker | -0.54 | -0.86 | -0.22 | -0.08 | <.01** |
| Observations | 1,558 |  |  |  |  |
| Adjusted R-squared | 0.01 |  |  |  |  |

*** p<0.001, ** p<0.01, * p<0.05

Supplemental Table 42: Adjusted Regression Model Examining Effect of Keyworker Status on Loneliness

|  | **Regression coefficient (B)** | **95% CI Lower** | **95% CI Upper** | ***β*** | ***p*** |
| --- | --- | --- | --- | --- | --- |
| Age (per decade) | -0.47 | -0.57 | -0.36 | -0.20 | <.001*** |
| Gender | -0.01 | -0.41 | 0.40 | -0.00 | 0.98 |
| Live alone | 2.58 | 2.18 | 2.98 | 0.30 | <.001*** |
| BAME Background | 0.31 | -0.13 | 0.75 | 0.03 | 0.16 |
| Risk Group |  |  |  |  |  |
| Most at Risk | 0.80 | 0.14 | 1.47 | 0.06 | 0.02* |
| Increased Risk | 0.41 | 0.03 | 0.78 | 0.05 | 0.03* |
| Health/Social Keyworker | -0.55 | -0.85 | -0.24 | -0.08 | <.001*** |
| Observations | 1,556 |  |  |  |  |
| Adjusted R-squared | 0.13 |  |  |  |  |

*** p<0.001, ** p<0.01, * p<0.05

Supplemental Table 43: Adjusted Regression Model Examining Interaction Between Keyworker Status and Gender on Loneliness

|  | **Regression coefficient (B)** | **95% CI Lower** | **95% CI Upper** | ***β*** | ***p*** |
| --- | --- | --- | --- | --- | --- |
| Age (per decade) | -0.47 | -0.58 | -0.36 | -0.20 | <.001*** |
| Gender | 0.37 | -0.37 | 1.10 | 0.04 | 0.32 |
| Live alone | 2.57 | 2.17 | 2.97 | 0.30 | <.001*** |
| BAME Background | 0.30 | -0.14 | 0.74 | 0.03 | 0.18 |
| Risk Group |  |  |  |  |  |
| Most at Risk | 0.80 | 0.13 | 1.46 | 0.06 | 0.02* |
| Increased Risk | 0.40 | 0.03 | 0.78 | 0.05 | 0.04* |
| Health/Social Keyworker | -0.62 | -0.95 | -0.29 | -0.10 | <.001*** |
| Gender#Health/Social Keyworker | 0.54 | -0.34 | 1.41 | 0.05 | 0.23 |
| Observations | 1,556 |  |  |  |  |
| Adjusted R-squared | 0.13 |  |  |  |  |

*** p<0.001, ** p<0.01, * p<0.05

Positive Mood

Supplemental Table 44: Unadjusted Regression Model Examining Effect of Keyworker Status on SPANE-PA Score

|  | **Regression coefficient (B)** | **95% CI Lower** | **95% CI Upper** | ***β*** | ***p*** |
| --- | --- | --- | --- | --- | --- |
| Health/Social Keyworker | 0.38 | -0.21 | 0.96 | 0.03 | 0.21 |
| Observations | 1,558 |  |  |  |  |
| Adjusted R-squared | 0.00 |  |  |  |  |

*** p<0.001, ** p<0.01, * p<0.05

Supplemental Table 45: Adjusted Regression Model Examining Effect of Keyworker Status on SPANE-PA Score

|  | **Regression coefficient (B)** | **95% CI Lower** | **95% CI Upper** | ***β*** | ***p*** |
| --- | --- | --- | --- | --- | --- |
| Age (per decade) | 0.39 | 0.18 | 0.60 | 0.09 | <.001*** |
| Gender | -0.65 | -1.44 | 0.13 | -0.04 | 0.10 |
| Live alone | -0.72 | -1.49 | 0.06 | -0.05 | 0.07 |
| BAME Background | -0.06 | -0.91 | 0.79 | -0.00 | 0.89 |
| Risk Group |  |  |  |  |  |
| Most at Risk | -0.97 | -2.26 | 0.32 | -0.04 | 0.14 |
| Increased Risk | -0.69 | -1.42 | 0.04 | -0.05 | 0.06 |
| Health/Social Keyworker | 0.39 | -0.20 | 0.97 | 0.03 | 0.20 |
| Observations | 1,556 |  |  |  |  |
| Adjusted R-squared | 0.01 |  |  |  |  |

*** p<0.001, ** p<0.01, * p<0.05

Supplemental Table 46: Adjusted Regression Model Examining Interaction Between Keyworker Status and Gender on SPANE-PA Score

|  | **Regression coefficient (B)** | **95% CI Lower** | **95% CI Upper** | ***β*** | ***p*** |
| --- | --- | --- | --- | --- | --- |
| Age (per decade) | 0.39 | 0.18 | 0.60 | 0.09 | <.001*** |
| Gender | -0.95 | -2.38 | 0.48 | -0.06 | 0.19 |
| Live alone | -0.71 | -1.49 | 0.07 | -0.05 | 0.07 |
| BAME Background | -0.05 | -0.90 | 0.80 | -0.00 | 0.91 |
| Risk Group |  |  |  |  |  |
| Most at Risk | -0.96 | -2.26 | 0.33 | -0.04 | 0.14 |
| Increased Risk | -0.69 | -1.41 | 0.04 | -0.05 | 0.07 |
| Health/Social Keyworker | 0.44 | -0.19 | 1.08 | 0.04 | 0.17 |
| Gender#Health/Social Keyworker | -0.42 | -2.13 | 1.29 | -0.02 | 0.63 |
| Observations | 1,556 |  |  |  |  |
| Adjusted R-squared | 0.01 |  |  |  |  |

*** p<0.001, ** p<0.01, * p<0.05

Perceived Risk from COVID-19

Supplemental Table 47: Unadjusted Regression Model Examining Effect of Keyworker Status on Perceived Risk from COVID-19

|  | **Regression coefficient (B)** | **95% CI Lower** | **95% CI Upper** | ***β*** | ***p*** |
| --- | --- | --- | --- | --- | --- |
| Health/Social Keyworker | 0.74 | 0.45 | 1.03 | 0.14 | <.001*** |
| Observations | 1,185 |  |  |  |  |
| Adjusted R-squared | 0.02 |  |  |  |  |

*** p<0.001, ** p<0.01, * p<0.05

Supplemental Table 48: Adjusted Regression Model Examining Effect of Keyworker Status on Perceived Risk from COVID-19

|  | **Regression coefficient (B)** | **95% CI Lower** | **95% CI Upper** | ***β*** | ***p*** |
| --- | --- | --- | --- | --- | --- |
| Age (per decade) | -0.06 | -0.17 | 0.04 | -0.04 | 0.22 |
| Gender | -0.08 | -0.48 | 0.32 | -0.01 | 0.69 |
| Live alone | -0.14 | -0.53 | 0.25 | -0.02 | 0.47 |
| BAME Background | 0.13 | -0.31 | 0.56 | 0.02 | 0.56 |
| Risk Group |  |  |  |  |  |
| Most at Risk | 0.15 | -0.49 | 0.79 | 0.01 | 0.65 |
| Increased Risk | 0.00 | -0.37 | 0.38 | 0.00 | 0.99 |
| Health/Social Keyworker | 0.75 | 0.46 | 1.05 | 0.15 | <.001*** |
| Observations | 1,184 |  |  |  |  |
| Adjusted R-squared | 0.02 |  |  |  |  |

*** p<0.001, ** p<0.01, * p<0.05

Supplemental Table 49: Adjusted Regression Model Examining Interaction Between Keyworker Status and Gender on Perceived Risk from COVID-19

|  | **Regression coefficient (B)** | **95% CI Lower** | **95% CI Upper** | ***β*** | ***p*** |
| --- | --- | --- | --- | --- | --- |
| Age (per decade) | -0.07 | -0.17 | 0.04 | -0.04 | 0.21 |
| Gender | 0.46 | -0.28 | 1.19 | 0.06 | 0.22 |
| Live alone | -0.15 | -0.53 | 0.24 | -0.02 | 0.46 |
| BAME Background | 0.12 | -0.32 | 0.55 | 0.02 | 0.60 |
| Risk Group |  |  |  |  |  |
| Most at Risk | 0.14 | -0.50 | 0.78 | 0.01 | 0.67 |
| Increased Risk | -0.00 | -0.38 | 0.37 | -0.00 | 0.98 |
| Health/Social Keyworker | 0.66 | 0.35 | 0.97 | 0.13 | <.001*** |
| Gender#Health/Social Keyworker | 0.77 | -0.11 | 1.65 | 0.09 | 0.09 |
| Observations | 1,184 |  |  |  |  |
| Adjusted R-squared | 0.02 |  |  |  |  |

*** p<0.001, ** p<0.01, * p<0.05
